# QUALITY OF LIFE AMONG INDIVIDUALS SEEKING FERTILITY TREATMENT IN UGANDA: FINDINGS FROM A CROSS-SECTIONAL FertiQoL SURVEY

**DOI:** 10.64898/2026.02.12.26346140

**Authors:** Daniel Zaake, Dinah Amongin, Anteneh Asefa, Mary Nakafeero, Christine K. Nalwadda, Anthony Kayiira, Suzanne N. Kiwanuka

**Author notes:** **Correspondence to** Daniel Zaake, Department of Health Policy Planning and Management, School of Public Health, College of Health Sciences, Makerere University, Department of Obstetrics and Gynecology, Mother Kevin Postgraduate School, Uganda Martyrs University, Nkozi, Kampala, Uganda.

## Abstract

**Background:** Infertility affects approximately one in six people globally and is associated with diminished quality of life (QoL). In Uganda, an estimated 6.4% of women experience infertility, yet evidence on fertility-specific QoL and its determinants remains limited. We assessed fertility-related QoL and associated factors among individuals seeking fertility care in public and private hospital settings in Kampala to inform patient-centred services.

**Methodology:** We conducted a hospital-based cross-sectional study (May 2024–January 2025) among 332 individuals aged 18–49 years attending fertility clinics at Kawempe National Referral Hospital (public; n=175) and St. Francis Hospital Nsambya (private; n=157). QoL was measured using the validated Core FertiQoL questionnaire, generating overall and domain scores (emotional, mind/body, relational, and social), transformed to a 0–100 scale (higher scores indicate better QoL). Mean scores were compared using independent-samples *t*-tests and one-way ANOVA. Multivariable linear regression identified factors independently associated with FertiQoL (α=0.05).

**Results:** Most participants were female (87.1%) and ≤40 years (88.0%). Secondary infertility was reported by 50.3%, and 58.4% had infertility duration <5 years. The overall mean FertiQoL score was 61.9 (SD 14.7), with similar totals across hospitals. Mind/body scores were comparatively higher, while emotional scores were lowest (mean 54.3, SD 21.3). In adjusted analyses, male gender (β=8.49; 95% CI 3.95–13.02; *p*<0.001) and secondary infertility (β=7.45; 95% CI 4.41–10.49; *p*<0.001) were associated with higher FertiQoL scores.

**Conclusions:** Fertility-related QoL among patients seeking care in Kampala was moderate and did not differ by facility type. Gender and infertility type were key correlates, underscoring the need for integrated, gender-sensitive psychosocial support alongside clinical infertility care.

## Background

Infertility constitutes a recognised public health issue, exerting a considerable influence on the quality of life of both men and women. Globally, approximately 48.5 million couples are affected by infertility, with the highest rates documented in South Asia, Sub-Saharan Africa, Northern Asia, and East/Central Europe (1). The experience of infertility is frequently distressing and may precipitate significant psychological and emotional repercussions, including hopelessness, guilt, shame, feelings of worthlessness, anxiety, depression, social withdrawal, and reduced self-esteem (2, 3).

According to the World Health Organization (WHO), quality of life (QoL) is defined as an individual’s perception of their position in life, considered within the context of their cultural and value systems and in relation to their aims, expectations, standards, and concerns (4). In recent years, QoL has emerged as a widely accepted endpoint in clinical research, reflecting a growing emphasis on patients’ experiences and preferences. When infertility treatment fails, individuals often experience frustration across social and psychological domains, as well as challenges to identity and life meaning (5). Those confronted with infertility face a range of personal, familial, and social challenges, navigating the complex interplay between their concerns and the coping strategies they adopt, which can have a significant impact on their QoL (6). The journey towards accepting infertility is multifaceted and shaped by various influences, including personality characteristics, individual aspirations, access to and affordability of healthcare, cultural beliefs, and the extent of familial and societal support available (7).

A systematic review using validated instruments—including the Fertility Quality of Life (FertiQoL) questionnaire, the Short Form Health Survey (SF-36), and the WHO Quality of Life (WHOQOL) tools—consistently shows that individuals experiencing infertility report poorer QoL than fertile counterparts (8). Evidence from sub-Saharan Africa similarly points to substantial psychosocial and relational burdens, yet fertility-specific QoL data remain limited and unevenly distributed across the region. In Uganda, qualitative findings by Asiimwe, Osingada (9) indicate that women experiencing infertility often face inadequate social support, marital discord, psychological distress and grief, unfulfilled expectations of motherhood, and significant financial strain (9). However, the extent to which these contextual stressors translate into measurable impairments in fertility-related QoL—and how such effects may differ by gender and care setting—has not been systematically quantified.

Assessing fertility-related QoL is important not only for documenting the lived burden of infertility at the individual level, but also for identifying relational impacts within families, stigma-related consequences at the community level, and potential gaps in the responsiveness and equity of infertility services within the health system. Such evidence can guide patient-centered counselling, inform psychosocial support integration, and support resource allocation and service planning, including the design of differentiated models of care across public and private facilities. Therefore, this study aims to assess fertility-related QoL among both men and women attending fertility clinics in two hospitals in Kampala, Uganda, and to examine associated sociodemographic, clinical, treatment-related, and psychosocial factors.

## Methods

### Study design

We conducted a hospital-based cross-sectional study between 1^st^ May 2024 and 30^th^ January 2025 among individuals aged 18–49 years seeking infertility care.

### Study setting

The study was implemented in fertility clinics within the outpatient Obstetrics and Gynaecology departments at two Kampala-based hospitals: Kawempe National Referral Hospital (KNRH; public) and St. Francis Hospital Nsambya (SFHN; private). The facilities were purposively selected to enable comparison across public and private service contexts and because both are high-volume institutions that provide structured infertility services within Kampala. KNRH is located approximately 8 km from Kampala’s central business district, and SFHN approximately 5 km from the central business district.

Both sites provide outpatient assessment and medical management for infertility (primary-level infertility care) as well as more specialized diagnostic and treatment services delivered through specialist obstetrician–gynecologists (secondary-level infertility care). Primary-level care in this context refers to initial evaluation, counselling, and first-line investigations and management, while secondary-level care refers to specialist-led diagnostic work-up and advanced management beyond first-line services.

### Study population

Eligible participants were men and women aged 18–49 years attending the infertility/fertility clinics at KNRH and SFHN during the study period, who provided written informed consent. Participants were recruited on clinic days: Fridays at KNRH (one clinic day per week; ∼15–20 patients per clinic day) and Wednesdays and Fridays at SFHN (two clinic days per week; ∼15 patients per clinic day).

### Sample size determination

The required sample size was 327 participants. This sample size was determined a priori using a single-population parameter approach for cross-sectional studies, incorporating a 95% confidence level, a 5% margin of error, and allowance for non-response (10).

### Sampling frame and sampling procedure

Systematic random sampling was used. The sampling frame comprised the clinic attendance registers for KNRH and SFHN, from which the total clinic population during the study period was estimated at 2,180 clients across both facilities. A sampling interval was calculated as *k* = *N*/*n =*2180/327 ≈ 6. A random start of *r=*4 was selected, after which every 6^th^ eligible client presenting to the clinic was approached for enrolment until the target sample size was achieved. On each clinic day, consecutive attendees were screened for eligibility, and those sampled were interviewed after informed consent.

### Variables

Quality of life was assessed using the Core Fertility Quality of Life (FertiQoL) module, a fertility-specific, self-administered questionnaire developed and validated by Boivin, Takefman (11). The FertiQoL instrument comprises two components: a Core module (24 items) and an optional Treatment module (10 items). In this study, only the Core module was administered to ensure relevance across participants irrespective of treatment status. Notably, we have translated the FertiQoL questionnaire into Luganda (12), the most widely spoken dialect in the study region, and validated it within the Ugandan context to ensure its linguistic and cultural appropriateness for the local population (13).

The Core FertiQoL evaluates fertility-related QoL across four domains: Emotional, Mind/Body, Relational, and Social. The Emotional domain captures infertility-related affective responses (e.g., sadness, grief, resentment) (11). The Mind/Body domain reflects perceived effects on physical functioning, cognition, and daily activities. The Relational domain assesses the impact of infertility on intimate partnership (e.g., communication, sexuality, commitment), while the Social domain captures effects on social inclusion and perceived expectations, stigma, and support (11).

Items are rated on 5-point response scales that are scored and, where applicable, reverse-coded so that higher values reflect better QoL. Domain and overall scores are computed and transformed to a 0–100 scale, with higher scores indicating better fertility-related QoL (11).

The scaled FertiQoL was the dependent variable ranging from 0 to 100. Independent variables were categorized as socio-demographic and fertility-related factors. The socio-demographic factors included marital status categorized as single, divorced, widow, cohabiting, or married; Age categorized as ≤40 years or >40 years; Educational level was categorized as no formal education, primary, secondary, and tertiary; Occupation categorized as employed and not employed; religion categorized as Catholic, Anglican, Muslim, and others. The fertility-related factors included type of infertility categorized as primary or secondary; Duration of infertility measured in years, and the health facility where the participant attended categorized as KNRH and SFNH.

### Data Collection

The administration of the self-reported FertiQoL questionnaire was overseen by the Principal Investigator (PI) with the assistance of two trained research assistants (RAs). Prior to the commencement of the pilot and main data collection phases, both RAs underwent structured training sessions to ensure standardized administration procedures and to enhance their familiarity with the instrument and ethical considerations. Eligible participants were provided with the FertiQoL Core module, which was self-administered in either English or Luganda, depending on participant preference, to facilitate comprehension and optimize response accuracy. The RAs were available to clarify any items and provide guidance while maintaining a non-intrusive presence to minimize social desirability bias. Responses were directly entered electronically via Kobo Toolbox (14), thereby reducing data entry errors and expediting data capture. Upon completion, all responses were systematically uploaded to a secure database. Comprehensive quality assurance protocols were implemented throughout the process, including real-time review of questionnaire completeness and consistency checks. Additionally, stringent measures were enacted to ensure participant anonymity and confidentiality, in accordance with ethical standards for cross-sectional survey research.

### Data Analysis

Data were analyzed using STATA Version 18 software (15). Categorical variables were summarized using frequencies and proportions. Fertility-related quality of life (QoL) scores were summarized using means and standard deviations.

Multicollinearity among candidate predictors was assessed using variance inflation factors (VIFs); a conservative threshold of VIF ≥3 was used to indicate potentially problematic collinearity, and no predictor exceeded this cut-off. To account for potential clustering by study site, we initially considered a mixed-effects linear regression model with study site as the clustering unit and a random intercept for site, while treating participant-level covariates as fixed effects. However, the intra-cluster correlation coefficient (ICC) for QoL by site was <0.001, indicating negligible between-site variance; therefore, a single-level linear regression framework was retained for subsequent association analyses.

Bivariable regression models were fitted to examine associations between QoL and sociodemographic and fertility-related characteristics. Variables with p <0.20 in bivariable analyses were carried forward into a multivariable linear regression model to estimate adjusted associations and minimize residual confounding. Model assumptions and fit were evaluated using standard diagnostic procedures, including assessment of linearity, normality of residuals, homoscedasticity, and influence/outliers.

Results are presented as adjusted β coefficients with 95% confidence intervals, and statistical significance was defined as p<0.05.

## Results

### Socio-Demographic and Fertility Characteristics of the Participants

Of the 332 participants (175 from KNRH and 157 from SFHN), the majority were 40 years or younger (292, 88.0%). Female participants (289, 87.1%) were more compared to the males (43, 12.9%). Nearly one-third (96, 28.9%) had primary or no formal education, while tertiary education was more common at SFHN (76, 48.4%). The most frequent occupations were in service and sales (88, 26.5%), followed by professional roles. The largest religious group was Catholic (98, 29.5%). Regarding infertility, half of the participants had secondary fertility (167, 50.3%), and most participants (195, 58.4%) reported having experienced infertility for less than five years *(Table 1)*.

**Table 1:** Socio-demographic and fertility characteristics of the participants.

|  | KNRH | SFHN | Total |
| --- | --- | --- | --- |
|  | n=175 | n=157 | n=332 |

| CHARACTERISTIC | Freq(col%) | Freq(col%) | Freq(col%) |
| --- | --- | --- | --- |
| **Age** |  |  |  |
| 40years or younger | 161(92.0) | 131(83.4) | 292(88.0) |
| Older than 40 years | 14(8.0) | 26(16.6) | 40(12.0) |
| **Gender** |  |  |  |
| Female | 154(88.0) | 135(85.9) | 289(87.1) |
| Male | 21(12.0) | 22(14.0) | 43(12.9) |
| **Marital status** |  |  |  |
| Married or domestic partnership | 175(100) | 157(100) | 332(100) |
| **Education level** |  |  |  |
| Primary or lower/No school | 60(34.2) | 36(22.9) | 96(28.9) |
| Secondary | 72(41.1) | 45(28.7) | 117(35.2) |
| Tertiary | 43(24.6) | 76(48.4) | 119(35.8) |
| **Occupation** |  |  |  |
| Professional | 23(13.1) | 38(24.2) | 61(18.4) |
| Technician and or associate professional | 18(10.3) | 27(17.2) | 45(13.6) |
| Service and sales worker | 44(25.1) | 44(28.0) | 88(26.5) |
| Others | 57(32.6) | 18(11.5) | 75(22.6) |
| Unemployed | 11(6.3) | 31(19.8) | 42(12.6) |
| **Religion** |  |  |  |
| Anglican | 41(23.4) | 32(20.4) | 73(22.0) |
| Catholic | 49(28.0) | 49(31.2) | 98(29.5) |
| Evangelical or Pentecostal | 35(20.0) | 17(10.8) | 52(15.7) |
| Moslem | 39(22.3) | 28(17.8) | 67(20.2) |
| Other religion | 11(6.3) | 31(19.8) | 42(12.6) |
| **Fertility type** |  |  |  |
| Primary infertility | 77(44.0) | 88(56.1) | 165(49.7) |
| Secondary infertility | 98(56.0) | 69(43.9) | 167(50.3) |
| **Duration of infertility** |  |  |  |
| Less than 5 years | 93(53.1) | 101(64.3) | 194(58.4) |
| 5 or more years | 82(46.9) | 56(35.7) | 138(41.6) |

### Quality of Life of Individuals Seeking Fertility Care

The overall mean FertiQoL score was 61.9 (SD = 14.7), with relatively similar totals across KNRH (61.8) and SFHN (61.9). Among the subscales, the relational domain had the highest internal consistency (α = 0.83) and a higher mean score at SFHN (64.9) compared to KNRH (59.6). Overall, mind-body scores were relatively high (68.2, SD = 20.7), while emotional well-being was lower (54.3, SD = 21.3). Social scores were comparable across sites (62.8, SD = 18.9). Reliability across the subscales ranged from α = 0.63 to 0.83, indicating acceptable internal consistency (***Error! Reference source not found***.).

### Average Quality of Life of The Participants by their Characteristics

Younger individuals (≤40 years, 292; 88.0%) had slightly lower scores (61.4) compared to those >40 years (65.3). Education showed a positive gradient, with tertiary-level participants (119; 35.8%) reporting the highest scores (63.8) compared to those with primary or no schooling (96; 28.9%, mean 59.5). By occupation, professionals (61; 18.4%) had the highest scores (64.4), while service and sales workers (88; 26.5%) reported the lowest (59.9). Religious differences were also noted, with Anglicans (73; 22.0%) and those in other religions (42; 12.6%) scoring higher (65.9 and 66.3, respectively), while Catholics (98; 29.5%) had the lowest (59.9). Participants with secondary infertility (167; 50.3%) had higher scores (65.2) than those with primary infertility (165; 49.7%, mean 58.5). Finally, those with infertility lasting less than five years (194; 58.4%) scored slightly higher (62.1) than those with infertility for five or more years (61.5) (**Error! Reference source not found**.).

### Factors associated with quality of life

Gender and fertility type were independently associated with the fertility quality of life. Male participants had significantly higher QoL scores than female participants (β = 8.49, 95% CI: 3.95– 13.02, p < 0.001). Participants with secondary infertility reported higher QoL scores compared to those with primary infertility (β = 7.45, 95% CI 4.41–10.49, p < 0.001). Catholic participants had lower fertility QoL scores compared to Anglicans; however, the association was not statistically significant (adjusted p = 0.008) *(Table 4)*.

**Table 2:** Fertility scores according to the different domains.

| Sub-scale | KNRH<br>Mean (SD) | SFHN<br>Mean (SD) | Overall<br>Mean (SD) | Cronbach reliability<br>coefficient($\alpha$) |
| --- | --- | --- | --- | --- |
| Emotional | 55.2(21.9) | 53.2(20.5) | 54.3(21.3) | 0.63 |
| Mind-Body | 69.2(21.9) | 67.1(19.3) | 68.2(20.7) | 0.63 |
| Relational | 59.6(14.2) | 64.9(18.1) | 62.2(16.3) | 0.83 |
| Social | 63.2(19.4) | 62.4(18.5) | 62.8(18.9) | 0.64 |
| Total | 61.8(14.8) | 61.9(14.7) | 61.9(14.7) | 0.76 |

**Table 3:**
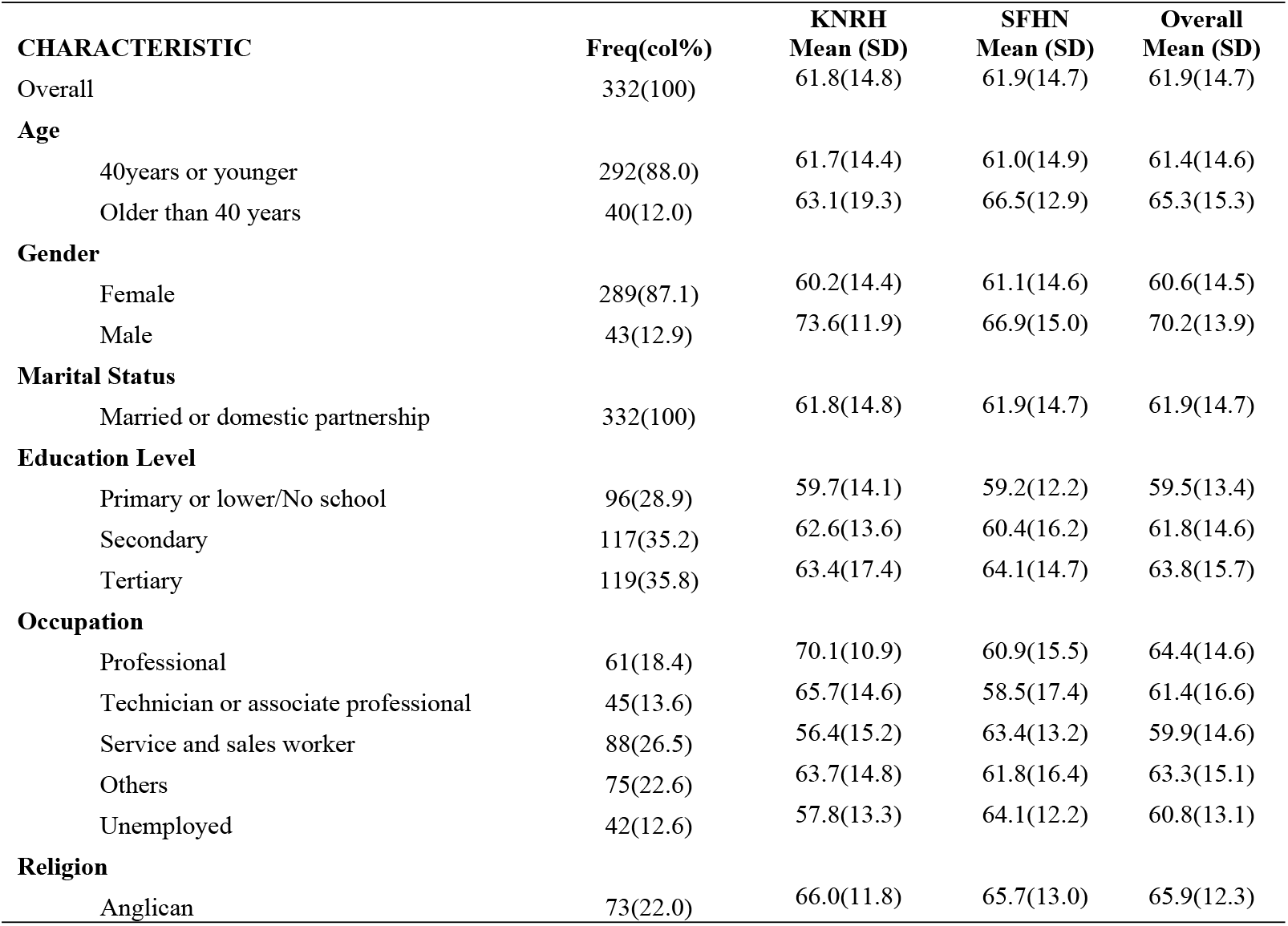
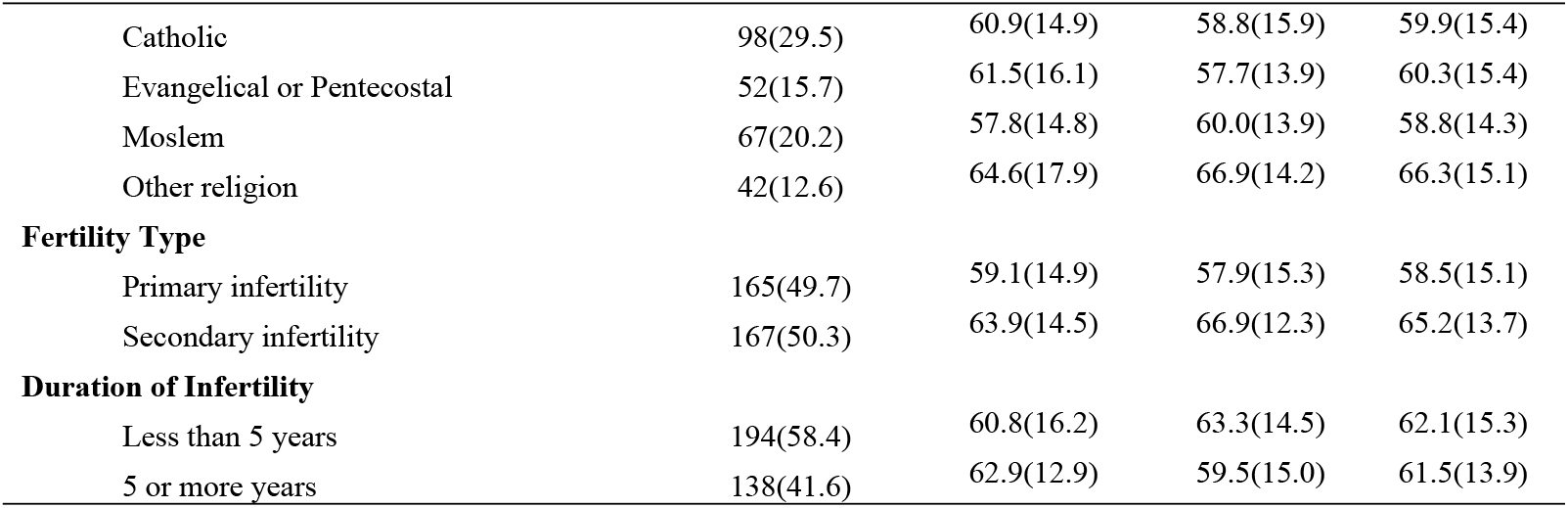
Average quality of life of the participants by participant characteristics.

| CHARACTERISTIC | Freq(col%) | KNRH<br>Mean (SD) | SFHN<br>Mean (SD) | Overall<br>Mean (SD) |
| --- | --- | --- | --- | --- |
| Overall | 332(100) | 61.8(14.8) | 61.9(14.7) | 61.9(14.7) |
| **Age** |  |  |  |  |
| 40years or younger | 292(88.0) | 61.7(14.4) | 61.0(14.9) | 61.4(14.6) |
| Older than 40 years | 40(12.0) | 63.1(19.3) | 66.5(12.9) | 65.3(15.3) |
| **Gender** |  |  |  |  |
| Female | 289(87.1) | 60.2(14.4) | 61.1(14.6) | 60.6(14.5) |
| Male | 43(12.9) | 73.6(11.9) | 66.9(15.0) | 70.2(13.9) |
| **Marital Status** |  |  |  |  |
| Married or domestic partnership | 332(100) | 61.8(14.8) | 61.9(14.7) | 61.9(14.7) |
| **Education Level** |  |  |  |  |
| Primary or lower/No school | 96(28.9) | 59.7(14.1) | 59.2(12.2) | 59.5(13.4) |
| Secondary | 117(35.2) | 62.6(13.6) | 60.4(16.2) | 61.8(14.6) |
| Tertiary | 119(35.8) | 63.4(17.4) | 64.1(14.7) | 63.8(15.7) |
| **Occupation** |  |  |  |  |
| Professional | 61(18.4) | 70.1(10.9) | 60.9(15.5) | 64.4(14.6) |
| Technician or associate professional | 45(13.6) | 65.7(14.6) | 58.5(17.4) | 61.4(16.6) |
| Service and sales worker | 88(26.5) | 56.4(15.2) | 63.4(13.2) | 59.9(14.6) |
| Others | 75(22.6) | 63.7(14.8) | 61.8(16.4) | 63.3(15.1) |
| Unemployed | 42(12.6) | 57.8(13.3) | 64.1(12.2) | 60.8(13.1) |
| **Religion** |  |  |  |  |
| Anglican | 73(22.0) | 66.0(11.8) | 65.7(13.0) | 65.9(12.3) |
| Catholic | 98(29.5) | 60.9(14.9) | 58.8(15.9) | 59.9(15.4) |
| Evangelical or Pentecostal | 52(15.7) | 61.5(16.1) | 57.7(13.9) | 60.3(15.4) |
| Moslem | 67(20.2) | 57.8(14.8) | 60.0(13.9) | 58.8(14.3) |
| Other religion | 42(12.6) | 64.6(17.9) | 66.9(14.2) | 66.3(15.1) |
| **Fertility Type** |  |  |  |  |
| Primary infertility | 165(49.7) | 59.1(14.9) | 57.9(15.3) | 58.5(15.1) |
| Secondary infertility | 167(50.3) | 63.9(14.5) | 66.9(12.3) | 65.2(13.7) |
| **Duration of Infertility** |  |  |  |  |
| Less than 5 years | 194(58.4) | 60.8(16.2) | 63.3(14.5) | 62.1(15.3) |
| 5 or more years | 138(41.6) | 62.9(12.9) | 59.5(15.0) | 61.5(13.9) |

**Table 4:** Linear Regression Analysis for Factors Associated with Quality of Life.

| CHARACTERISTIC | Crude coefficient (95% CI) | p-value | Adjusted coefficient (95% CI) | p-value |
| --- | --- | --- | --- | --- |
| **Study site** |  |  |  |  |
| KNRH | Ref |  |  |  |
| SFHN | 0.11(-3.08,3.30) | 0.945 | - |  |
| **Age** |  |  |  |  |
| 40 years or younger | Ref |  | Ref |  |
| Older than 40 years | 3.89(-0.98,8.77) | 0.117 | 0.85(-3.86,5.57) | 0.722 |
| **Gender** |  |  |  |  |
| Female | Ref |  | Ref |  |
| Male | 9.59(4.96,14.22) | <0.001 | 8.49(3.95,13.02) | <0.001 |
| **Education level** |  |  |  |  |
| Primary or lower/No school | Ref |  | Ref |  |
| Secondary | 2.24(-1.73,6.21) | 0.268 | 2.89(-0.88,6.67) | 0.113 |
| Tertiary | 4.29(0.35,8.25) | 0.034 | 3.26(0.64,7.16) | 0.101 |
| **Occupation** |  |  |  |  |
| Professional | Ref |  |  |  |
| Technician or associate professional | -3.03(-8.72,2.66) | 0.327 | - |  |
| Service and sales worker | -4.48(-9.30,0.35) | 0.066 | - |  |
| Others | -1.13(-6.13,3.86) | 0.658 | - |  |
| Unemployed | -3.62(-8.82,1.57) | 0.147 | - |  |
| **Religion** |  |  |  |  |
| Anglican | Ref |  | Ref |  |
| Catholic | -5.98(-10.15, -1.83) | 0.005 | -5.69(-9.91, -1.49) | 0.008 |
| Evangelical or Pentecostal | -5.62(10.68, -0.56) | 0.030 | -4.58(-9.52,0.36) | 0.069 |
| Moslem | -7.14(-11.59, -2.69) | 0.002 | -5.46(-10.09, -0.83) | **0.021** |
| Other religion | 0.39(-4.96,5.76) | 0.884 | 1.29(-4.04,6.62) | 0.634 |
| **Fertility type** |  |  |  |  |
| Primary infertility | Ref |  | Ref |  |
| Secondary infertility | 6.72(3.61,9.82) | <0.001 | 7.45(4.41,10.49) | **<0.001** |
| **Duration of infertility** |  |  |  |  |
| Less than 5 years | Ref |  | - |  |
| 5 or more years | -0.54(-3.72,2.64) | 0.738 | - |  |

## Discussion

The overall FertiQoL score in this study was moderate, with an average of 61.9 (SD 14.7). This score was almost the same in both public (KNRH: 61.8) and private (SFHN: 61.9) hospitals. This suggests that the type of hospital does not significantly affect the QoL of women experiencing infertility (16). Similar FertiQoL scores have been found in India and China, where women’s mean QoL score was 66.1 and 64.5, respectively (16, 17). In contrast, a study from Tanzania reported a higher average score of 70.6 (8). The contrast between Tanzania and Uganda may be due to the higher education levels found in Tanzania, which are often associated with better awareness of infertility treatments, greater access to healthcare resources, and more effective coping strategies, all of which can contribute to higher fertility-related QoL scores (8).

The relational domain had the highest internal consistency (α=0.83) and a higher average score at SFHN (64.9) compared to KNRH (59.6). This means that individuals in the private setting experienced better relational well-being, possibly because SFHN serves individuals with better resources and access to counselling, leading to stronger family or spousal support. Similar relational scores have been reported internationally. For instance, Chinese IVF patients had an average of 67.9 (16). On the other hand, Pakistani women had much lower relational scores of 47.3, and Tanzanian women reported 50.8, both below our findings (8, 18). These differences may be explained by cultural and structural factors. In Pakistan and some African contexts, infertility is strongly stigmatized and frequently attributed to women, often causing marital strain and polygamy, whereas in Uganda, the stigma is less severe, leading to comparatively lower relational strain (19-21).

The mind-body subscale scores in our study were relatively high, with an average of 68.2 (SD 20.7), suggesting that individuals generally maintained good physical and mental health despite dealing with infertility. Similar findings were observed in India, where non-obese women reported higher mind-body scores (17). In contrast, Chinese and Tanzanian women had lower scores, averaging about 63.4 and 59.5, respectively, while Pakistani women had even lower scores at around 50.7, probably reflecting higher health burdens or stress (16-18). The relatively high mind-body scores in Uganda suggest that the impact of infertility is felt more in emotional and social ways than in physical health, unlike situations where obesity or serious health problems worsen physical and mental stress.

On the other hand, emotional well-being scores were lower, with an average of 54.3 (SD 21.3), indicating that infertility caused the most significant emotional strain for individuals. The lower emotional QoL observed may relate to strong cultural pressures to have children in Uganda, along with fears of lifelong childlessness and limited access to psychological help (19). This finding aligns with studies in India, where the emotional aspect was notably affected, with women feeling a lot of anxiety and sadness (17). Additionally, a study on Pakistani women had emotional scores similar to the ones in our study, averaging around 53.3. (18) In contrast, a study on Chinese women reported slightly higher emotional scores at 64.1, while Tanzanian women had very high scores at 77.4, possibly due to stronger emotional support networks, higher education levels that facilitate coping with infertility-related stress, and, in the case of China, the advantages of being a more developed country with greater resources and healthcare infrastructure, which together enhance the overall quality of care and emotional well-being of patients(8, 16).

The average social domain scores were 62.8 (SD 18.9) and showed no difference between KNRH and SFHN. This suggests that the social challenges of infertility, such as stigma and community interactions, are similar across care settings. This consistency likely stems from shared cultural expectations around fertility in Uganda, which do not significantly differ between women seeking care at public and private facilities (19). Globally, social scores were higher in China (69.9) and Tanzania (68.9), while Pakistani women scored lower at 57.4 (16, 18, 22). These differences show that individuals in Uganda face moderate social challenges, which are less severe than those in countries with strong stigma, like Pakistan, but not as favourable as in places where higher education and strong community support lessen the social impact of infertility (19-22).

Finally, the internal consistency of the FertiQoL subscales ranged from Cronbach’s α=0.63 to 0.83, indicating acceptable reliability. The relational subscale (α=0.83) was strongest, while other scales (e.g., environment, with the lowest α≈0.63) were above the conventional acceptability threshold. This mirrors the original FertiQoL validation, which reported Cronbach α values from about 0.72 up to 0.92 across its scales (2). It also aligns with the Tanzanian study’s report of alphas 0.64-0.9 (2, 22). Thus, despite cultural differences, the FertiQoL instrument retained cohesive structure in our Ugandan sample. The robust α for relational echoes other findings (e.g., the Chinese version has α≈0.92), suggesting that questions about partner/family support form a tightly related construct (2, 22). The somewhat lower α for other domains is not unusual and is comparable to previous cross-cultural work (2, 22). Overall, the acceptable reliability supports the validity of our findings across domains.

Regarding factors associated, this study found that gender and fertility type were associated with fertility-related QoL. Consistent with prior research, men reported better QoL than women. In many sociocultural contexts, fertility challenges place an emotional and social burden on women, who are always held primarily responsible for childbearing. Such expectations expose women to stigma, blame, and familial or community pressure (23), which in turn contribute to heightened stress, anxiety, and diminished well-being (24). More recent evidence reinforces this pattern: Mahfoudh, Braham (25) revealed that Tunisian women reported lower relational and emotional QoL compared to men (25), while a cross-sectional study in Europe found similar gender disparities in fertility-related QoL outcomes (26). Likewise, Cusatis, Fergestrom (27) highlighted that women’s QoL was more negatively impacted by the time and emotional demands of treatment (27). These findings indicate that gendered experiences of infertility contribute to persistent disparities in QoL. Addressing these inequities requires gender-sensitive psychosocial interventions and broader health system strategies that acknowledge the cultural and relational dimensions of infertility.

This study also found that participants with secondary infertility reported better QoL scores than those with primary infertility. Having at least one child appears to provide a degree of social reassurance and reduce external pressure, even when couples later face fertility difficulties (9). By contrast, individuals with primary infertility encounter more intense social scrutiny, fear of stigma, and emotional distress, which can negatively affect daily functioning and well-being (24, 28). In our sample, participants with secondary infertility scored on average 7.4 points higher than those with primary infertility, a difference that is consistent with other findings. Research from India (29), China (30), and Turkey QoL (31) has shown that women with primary infertility report poorer QoL outcomes than those with secondary infertility, with Turkish studies further noting that prior childbearing reduces depression and enhances QoL (31). This global trend suggests that prior pregnancy may confer psychological benefits in the face of later infertility, possibly by reducing blame or demonstrating fertility potential.

However, not all contexts show the same pattern. For example, a Zanzibar study reported lower QoL among women with secondary infertility compared to those with primary infertility, showing how cultural expectations for larger families may invert the usual association (22). Such contrasts indicate that while the majority of evidence supports better QoL among secondary-infertile patients, local cultural norms and health system factors can shape these experiences in various ways. These findings emphasize the need for culturally sensitive psychosocial interventions that account for both the protective effects of prior childbearing and the unique pressures faced in different settings.

Other sociodemographic factors, including age, education level, occupation, religion, and duration of infertility, did not show statistically significant associations with fertility-related QoL in our dataset. This contrasts with findings from some studies in other contexts. For example, recent studies in Europe (26) and Nigeria (32) reported that longer duration of infertility and older age were associated with poorer QoL outcomes (26, 32). Similarly, studies in Cameroon (33) and Tunisia (25) found that higher education was associated with better coping and improved QoL (25, 33). These results highlight the importance of examining infertility experiences across diverse populations, as predictors of QoL may vary depending on social norms, health system structures, and community expectations.

Despite these limitations, a key strength of this study is the direct comparison between a public and a private healthcare facility, providing unique insights into how care setting influences different domains of quality of life. The use of the validated FertiQoL instrument ensures the reliability of our outcome measures and allows for international comparability. The study also identifies under-researched predictors of QoL, such as religious affiliation, within the Sub-Saharan African context. Additionally, the substantially large sample size enhances the robustness and generalizability of the findings across diverse patient populations.

## Conclusion

Infertility is not only a medical condition but a deeply social and emotional experience, shaped by cultural expectations, gender roles, and personal circumstances. The patterns observed in this study reinforce the importance of viewing fertility challenges through a broader lens that accounts for stigma, relational strain, and psychosocial well-being. While some factors may vary across contexts, the consistent message is that infertility care must extend beyond clinical treatment to include compassionate, gender-sensitive, and culturally responsive support. By integrating medical, psychological, and social dimensions, health systems can better address the complex realities of infertility and improve the overall quality of life for those affected.

## Data Availability

The data underlying the results presented in the study are available from: Dr Daniel Zaake (corresponding author) Department of Health Policy Planning and Management, School of Public Health, College of Health Sciences, Makerere University

## List of abbreviations

ANOVA: Analysis of Variance
ASRM: American Society of Reproductive Medicine
CI: Confidence Interval
ESHRE: European Society of Human Reproduction and Embryology
FertiQoL: Fertility Quality of Life
IVF: In Vitro Fertilization
KNRH: Kawempe National Referral Hospital
QoL: Quality of Life
SD: Standard Deviation
SFHN: St. Francis Hospital Nsambya
VIF: Variance Inflation Factor
WHO: World Health Organization

## Declarations

### Ethics approval and consent to participate

Ethical approval was obtained from the Ministry of Health of Uganda to cover facilities nationwide. Additional approval was secured from the Uganda National Council for Science and Technology and the Makerere University School of Public Health Research and Ethics Committee (REC Protocol Number: SPH 2023-525). Administrative permission was also obtained from all the health facilities where the study was conducted. Written informed consent was obtained from all respondents before the start of data collection.

**Consent for publication:** Not applicable

**Availability of data and materials:** Data is available upon request. Requests should be sent to

**Conflict of interest:** None to declare.

**Funding/Sponsorship:** Self-sponsored.

### Authors’ contributions

DZ conceived and designed the study, secured necessary approvals, and oversaw the study’s execution. Additionally, DZ conducted the literature review, performed the data analysis, interpreted the results, prepared the tables and figures, and drafted and finalized the manuscript. DA, CKN, MN, AK, AA, and SNK contributed to the study design, data analysis, interpretation of the results, and provided substantial revisions to the manuscript. All co-authors participated in the redrafting and finalization of the article.

## References

1. Sripad P, Desai S, Regules R, Chakraborty S, Habib H, Viloria A, et al. Exploring experiences of infertility amongst women and men in low-income and middle-income countries: protocol for a qualitative systematic review. BMJ Open. 2021;11:e050528.

2. Boivin J, Takefman J, Braverman A. The Fertility Quality of Life (FertiQoL) tool: development and general psychometric properties. Fertil Steril. 2011;96(2):409–15.e3.

3. Bueno-Sánchez L, Alhambra-Borrás T, Bonilla-Campos A, Durá-Ferrandis E. Psychometric characteristics of core FertiQoL questionnaire in a Spanish sample of infertile women. Anales de Psicología/Annals of Psychology. 2024;40(3):364–72.

4. The World Health Organization quality of life assessment (WHOQOL): Position paper from the World Health Organization. Social Science & Medicine. 1995;41(10):1403–9.

5. Wdowiak A, Anusiewicz A, Bakalczuk G, Raczkiewicz D, Janczyk P, Makara-Studzińska M. Assessment of Quality of Life in Infertility Treated Women in Poland. International Journal of Environmental Research and Public Health. 2021;18(8).

6. Kiani Z, Simbar M, Hajian S, Zayeri F. Quality of life among infertile women living in a paradox of concerns and dealing strategies: A qualitative study. Nurs Open. 2021;8(1):251–61.

7. Obiegło M, Uchmanowicz I, Wleklik M, Jankowska-Polańska B, Kuśmierz M. The effect of acceptance of illness on the quality of life in patients with chronic heart failure. Eur J Cardiovasc Nurs. 2016;15(4):241–7.

8. Suleiman M, August F, Nanyaro MW, Wangwe P, Kikula A, Balandya B, et al. Quality of life and associated factors among infertile women attending infertility clinic at Mnazi Mmoja Hospital, Zanzibar. BMC Women’s Health. 2023;23(1):400.

9. Asiimwe S, Osingada CP, Mbalinda SN, Muyingo M, Ayebare E, Namutebi M, et al. Women’s experiences of living with involuntary childlessness in Uganda: a qualitative phenomenological study. BMC Women’s Health. 2022;22(1):532.

10. Lwanga SK, Lemeshow S. Sample size determination in health studies: a practical manual. Geneva: World Health Organization; 1991.

11. Boivin J, Takefman J, Braverman A. The fertility quality of life (FertiQoL) tool: development and general psychometric properties. Hum Reprod. 2011;26(8):2084–91.

12. Makumbi D, Kayiira A, Tumwine G, Byaruhanga RN, Zaake D. Translation and cultural adaptation of the FertiQoL questionnaire into Luganda: A comprehensive report. Afr J Reprod Health. 2025;29(5):29–35.

13. Makumbi D, Kayiira A, Tumwine G, Byaruhanga R, Kiwanuka S, Dina A, et al. Psychometric Evaluation of the Fertiqol Questionnaire Among Infertile Individuals and Couples in Uganda: Findings From a Tertiary Hospital in Peri-urban Kampala 2025.

14. KoBoToolbox. Introduction to KoboToolbox 2026 [updated 2026–01–27. Available from: https://support.kobotoolbox.org/welcome.html.

15. StataCorp. Stata Statistical Software: Release 18. College Station, TX: StataCorp LLC; 2023.

16. Song D, Li X, Yang M, Wang N, Zhao Y, Diao S, et al. Fertility quality of life (FertiQoL) among Chinese women undergoing frozen embryo transfer. BMC Women’s Health. 2021;21(1):177.

17. Desai HJ, Gundabattula SR. Quality of life in Indian women with fertility problems as assessed by the FertiQoL questionnaire: a single center cross sectional study. J Psychosom Obstet Gynaecol. 2019;40(1):82–7.

18. Abbasi S, Kousar R. The fertility quality of life (FertiQol) questionnaire in Pakistani infertile women. Journal of Bahria University Medical and Dental College. 2016;6(3):170–3.

19. Asiimwe S, Osingada CP, Mbalinda SN, Muyingo M, Ayebare E, Namutebi M, et al. Women’s experiences of living with involuntary childlessness in Uganda: a qualitative phenomenological study. BMC Womens Health. 2022;22(1):532.

20. Tabassum A, Sadia R, Huda S, Khan S. Infertility-related Stress and Marital Satisfaction among Pakistani Infertile Individuals. 2023;5:71–81.

21. Lucas D, Mahiti G. “His biological mother told him; you do not have children”. The lived experience and coping strategies of married women living with infertility in Tanzania. East African Journal of Public Health. 2024;16(1).

22. Suleiman M, August F, Nanyaro MW, Wangwe P, Kikula A, Balandya B, et al. Quality of life and associated factors among infertile women attending infertility clinic at Mnazi Mmoja Hospital, Zanzibar. BMC Womens Health. 2023;23(1):400.

23. Lo SS, Kok WM. Sexual functioning and quality of life of Hong Kong Chinese women with infertility problem. Hum Fertil (Camb). 2016;19(4):268–74.

24. Howe S, Zulu JM, Boivin J, Gerrits T. The social and cultural meanings of infertility for men and women in Zambia: legacy, family and divine intervention. Facts Views Vis Obgyn. 2020;12(3):185–93.

25. Mahfoudh K, Braham M, Askri F, Ouali U, Zgueb Y, Chakroun N, et al. Factors influencing quality of life in Tunisian infertile couples: a gender-based analysis. Middle East Fertility Society Journal. 2025;30(1):57.

26. Celda-Belinchón L, Saus-Ortega C, Palop-Muñoz J, Rubio Rubio JM, Ferrandis ED. Quality of life in women and men with infertility: A cross-sectional study. European Journal of Obstetrics and Gynecology and Reproductive Biology. 2025;313.

27. Cusatis R, Fergestrom N, Cooper A, Schoyer KD, Kruper A, Sandlow J, et al. Too much time? Time use and fertility-specific quality of life among men and women seeking specialty care for infertility. BMC Psychology. 2019;7(1):45.

28. Kuug AK, James S, Sihaam J-B. Exploring the cultural perspectives and implications of infertility among couples in the Talensi and Nabdam Districts of the upper east region of Ghana. Contraception and Reproductive Medicine. 2023;8(1):1–10.

29. Bose S, Roy B. Fertility related quality of life in primary infertile couples: a comparative study from Eastern India. The International Journal of Indian Psychology. 2017.

30. Shi Z, Zheng Y, Zhu X, Mao Z, Nie H, Chen G, et al. Understanding health-related quality of life in Chinese infertility patients: a qualitative study. Quality of Life Research. 2025.

31. Turgay B, Aytaç R. Investigation of depression frequency and quality of life in couples with infertility complaints in Turkey. Scientific Reports. 2025;15(1):26519.

32. Amadi L, Raji FO, Odelola OI. The Effects of Infertility on the Quality of Life of Women: A Comparative Study in a Nigerian Tertiary Centre. Nigerian Journal of Medicine. 2025;34(1):38–44.

33. Mengne G, Emenguele P, Kobenge F. Psychosocial Effects of Infertility among Infertile Women Attending the Outpatient Consultation Unit of Chracerh Yaoundé Cameroon; a Cross-Sectional Study. Gynecol Reprod Health. 2024; 8 (3): 1–9. Correspondence: Prof Thomas Obinchemti Egbe, MD, Department of Obstetrics and Gynecology, Faculty of Health Sciences, University of Buea, Phone:+ 237677450005 Received. 2024;14.

